# Encrypted federated learning for secure decentralized collaboration in cancer image analysis

**DOI:** 10.1101/2022.07.28.22277288

**Authors:** Daniel Truhn, Soroosh Tayebi Arasteh, Oliver Lester Saldanha, Gustav Müller-Franzes, Firas Khader, Philip Quirke, Nicholas P. West, Richard Gray, Gordon G. A. Hutchins, Jacqueline A. James, Maurice B. Loughrey, Manuel Salto-Tellez, Hermann Brenner, Alexander Brobeil, Tanwei Yuan, Jenny Chang-Claude, Michael Hoffmeister, Sebastian Foersch, Tianyu Han, Sebastian Keil, Maximilian Schulze-Hagen, Peter Isfort, Philipp Bruners, Georgios Kaissis, Christiane Kuhl, Sven Nebelung, Jakob Nikolas Kather

## Abstract

Artificial Intelligence (AI) has a multitude of applications in cancer research and oncology. However, the training of AI systems is impeded by the limited availability of large datasets due to data protection requirements and other regulatory obstacles. Federated and swarm learning represent possible solutions to this problem by collaboratively training AI models while avoiding data transfer. However, in these decentralized methods, weight updates are still transferred to the aggregation server for merging the models. This leaves the possibility for a breach of data privacy, for example by model inversion or membership inference attacks by untrusted servers. Homomorphically encrypted federated learning (HEFL) is a solution to this problem because only encrypted weights are transferred, and model updates are performed in the encrypted space. Here, we demonstrate the first successful implementation of HEFL in a range of clinically relevant tasks in cancer image analysis on multicentric datasets in radiology and histopathology. We show that HEFL enables the training of AI models which outperform locally trained models and perform on par with models which are centrally trained. In the future, HEFL can enable multiple institutions to co-train AI models without forsaking data governance and without ever transmitting any decryptable data to untrusted servers.

**One Sentence Summary:** Federated learning with homomorphic encryption enables multiple parties to securely co-train artificial intelligence models in pathology and radiology, reaching state-of-the-art performance with privacy guarantees.

## Introduction

Artificial intelligence (AI) and machine learning techniques are transforming cancer imaging and cancer research and will have a profound impact on the practice of medicine^1–4^. They can automate manual tasks in medical image analysis and can be used to extract hidden information from routinely available clinical image data, beyond what is visible to the human eye^5, 6^. AI models have been used for the detection and diagnosis of cancer, subtype classification, and optimization of cancer treatments. In particular, deep neural networks have been trained to analyze radiology images and digitized pathology slides for numerous different cancer types. For example, AI models can now detect mammographic lesions with expert-level performance^7^. Similarly, AI models predict molecular biomarkers for treatment selection directly from routine pathology slides of solid tumors^8–13^.

However, the training of AI models is infamously data hungry and requires large amounts of annotated training data. While this data may already exist, in most cases it is scattered among multiple centers. Collecting this data at a central site is hindered by obstacles which are often insurmountable in practice, most notably issues with data privacy and data governance. The data governance problem has been addressed by collaborative learning protocols such as federated learning (FL)^14, 15^ in which an AI model is trained on separate sites and in which not data, but only the learned model weights are shared. This facilitates collaboration between multiple parties, but still poses significant risks for breach of patient privacy. The weight updates communicated to the central FL server contain information about the data that can be extracted to reconstruct sensitive patient information^16^. This can be exploited through privacy attacks such as model inversion^17–19^, in which a malicious server eavesdropper captures the weight updates and attempts to recover the private dataset used to train the model or reveal other private attributes. One measure to protect against privacy breaches is Differential Privacy in which deliberate noise is added to the training updates by each site ^18^. However, while this paradigm protects private information, it comes at a utility tradeoff and can lead to less performant AI models as demonstrated recently ^20^. In our study we employ Homomorphic encryption (HE). HE can protect against a malicious server eavesdropper while maintaining AI model performance by encrypting the weight updates before sending them to the central server. The central server then performs the weight aggregation on the encrypted values and the encrypted updated weights are sent back to the clients for decryption and incorporation into their models. Importantly, since the central server does not have access to the decryption key, it cannot infer any information about which calculations have been done at individual peer locations and thus cannot extract sensitive private information. In other words, all handling of the model parameters happens in the encrypted space, making homomorphic encryption an optimal tool for low-trust environments and handling of personal health data.

In this study, we examined how HEFL can be leveraged for training of competitive AI models for cancer diagnosis and detection of cancer biomarkers in radiology and pathology images. To this end, we assumed the following threat model: A mutually trusting confederation of data owners wishes to collaboratively train a model on their joint data, but neither wants to relinquish data governance. For conducting the training, the confederation makes use of an untrusted aggregation server, which we assume to honestly participate in the protocol (i.e., faithfully conduct the aggregation procedure), but attempt to extract all available information from the weight updates sent to it by the other participants (honest but curious threat model). We evaluated the training of AI models in three retrospective multicentric settings: 1) AI models are trained with local data only 2) AI models are trained with conventional federated learning whereby no additional measure of protection against privacy-centred attacks on the updates is undertaken and 3) AI models are trained with HEFL in a decentralized, secure and privacy-preserving manner, whereby the individual participants encrypt their weight update before transmitting it to the server. We hypothesized that the collective and secure training of AI models reaches better accuracy than training of local models and is associated with minimal risk of privacy leakage as compared to conventional FL while keeping the cost of additional training time low.

## Results

### HEFL guarantees data privacy compared to conventional federated learning in the untrusted central server setting

When multiple institutions collaborate in a conventional federated learning scheme, weight updates are calculated locally and are sent to a central server to be aggregated. When unencrypted weight updates are transmitted, we demonstrate that the untrusted central server can reconstruct the training images from the weight updates in a model inversion attack. In this setting we train a neural network for the detection of malignant lesions on brain MRI examinations from the brain tumor segmentation (BraTS) dataset ^21–23^. We employ a realistic setting in which data is contributed by five different institutions and in which each institution performs separate weight updates only on their data. We then perform a gradient inversion attack following the approach by Zhao et al. ^24^. We demonstrate that the original training images can be reconstructed after only 120 iterations - notably, before training of the underlying neural network objective has converged, see Figure 1. This poses a serious threat and renders the whole concept of conventional federated learning vulnerable to privacy-focused attacks. To showcase that homomorphic encryption can be used to counter these attacks and to salvage patient privacy, we repeat the training procedure, but employ homomorphic encryption in which the central server only has access to the encrypted weight updates and the key is kept private by the peers. Following the same approach - no identifiable information can be extracted from the weight updates, even after eavesdropping on the weight updates for 40,000 iterations.

**Fig. 1:**
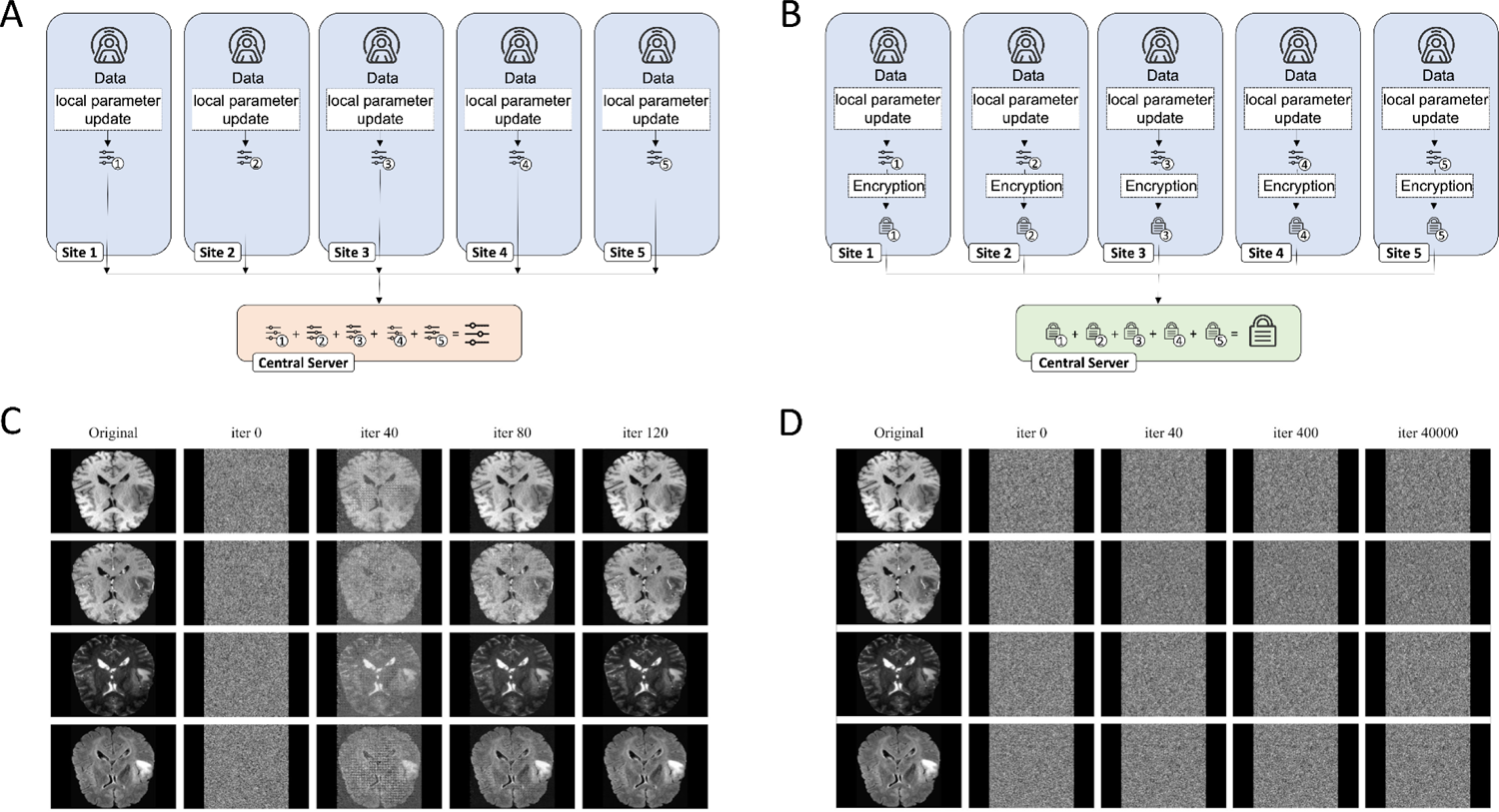
Schematic of FL and HEFL and associated Information extraction attacks. **(A)** In FL, each site trains on their own data and weight updates are transmitted to the central server for aggregation. **(B)** In HEFL, the weight updates are encrypted and the server only has access to the encrypted values. While FL allows the server to extract patient sensitive information by reconstructing the images from the weights through gradient inversion attacks and eavesdropping on the weight updates **(C)**, this information remains protected in HEFL and images can not be reconstructed **(D)**. Experiments were performed on 2D slices including native T1-weighted sequences in the top row, post-contrast T1-weighted sequences in the second row, T2-weighted sequences in the third row and fluid attenuated inversion recovery sequences in the bottom row.

### Secure training does not affect performance of oncological AI models

We trained AI models for tasks in oncology spanning both radiological and histopathological use-cases, see Figure 2. Each model was trained in three settings: a) AI models are trained with local data only b) AI models are trained with conventional federated learning in a decentralized manner c) AI models are trained with HEFL in a decentralized, secure and privacy-preserving manner. While approach a) is immune to privacy leaks, it results in training on only a limited subset of the possible data pool. Approach b) makes full use of the data but is prone to privacy leaks through the aforementioned attack by the untrusted aggregator. Only approach c) combines both training on full data and guarantees patient privacy. Moreover, as the HE scheme utilized in our study is endowed with a correctness guarantee (i.e. the values of the decrypted updates are guaranteed to be identical up to numerical precision to their plain-text counterparts), this setting does not suffer from an accuracy penalty compared to non-private training. We test the performance of each paradigm for AI models for the segmentation of glioblastoma on magnetic resonance images (MRI) and for the detection of microsatellite instability in histopathological whole slide images (WSI) of colorectal cancer patients.

**Fig. 2:**
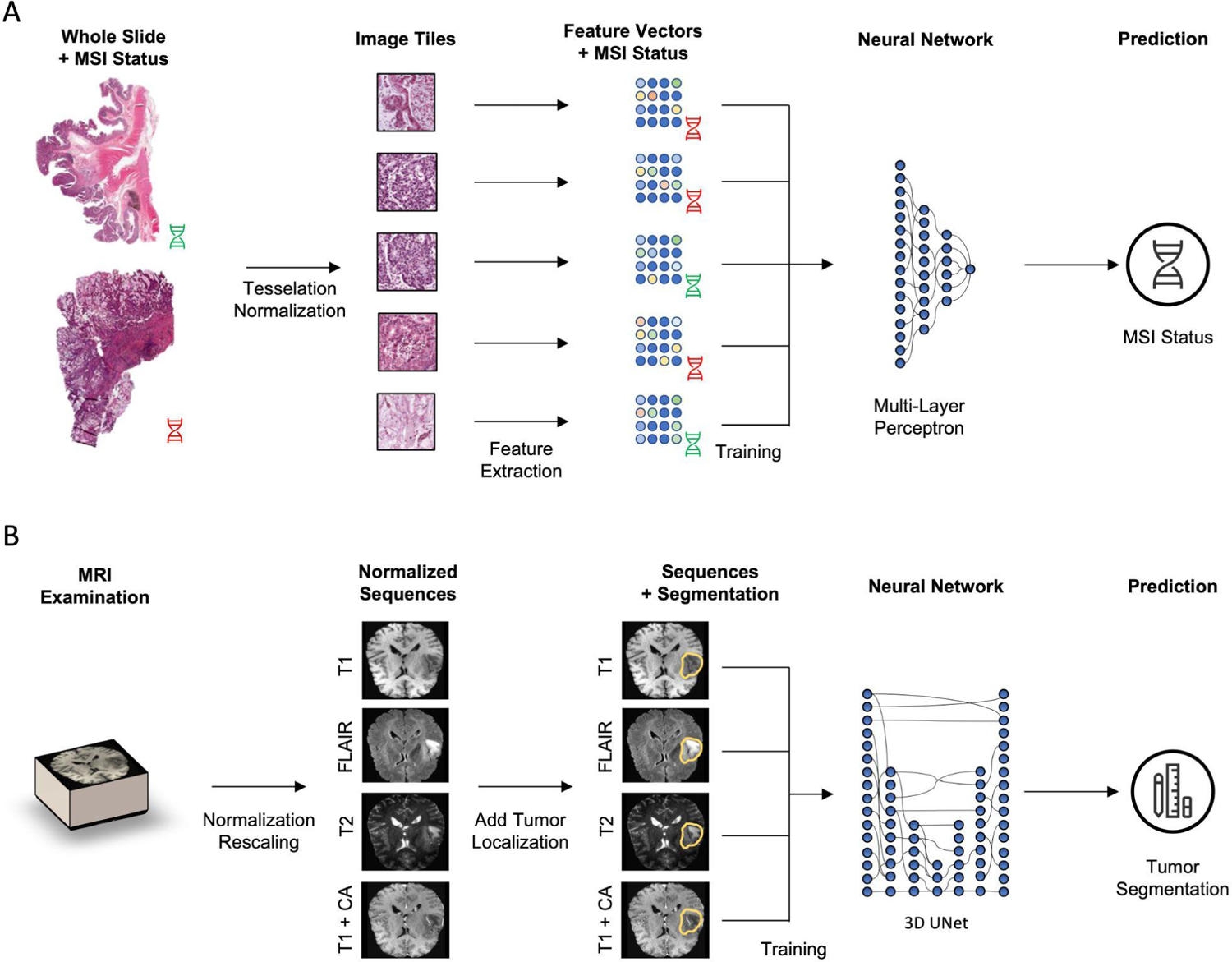
Schematic of the Deep Learning workflow. **(A)** Histology images are first tessellated. Features are then extracted by a feature extractor network (fixed) and a multi-layer perceptron is trained to predict MSI status. **(B)** The MRI examination is normalized and rescaled to a standard resolution of 128×128×128. All four three-dimensional sequences are then fed into a UNet architecture that is trained to predict tumor segmentation outlines.

### Segmentation of glioblastoma on MRI

The BraTS training dataset comprises 369 MRI examinations of 369 patients which have been acquired at seventeen different clinical centers. We partitioned the data along the information where the images had been acquired into five groups and trained a U-Net architecture to segment the tumor volume. All models were tested on an external test set from a separate institution provided by the BraTS organizers (n=125) and employed the dice similarity score as a measure of performance. All five locally trained AI models performed inferior in terms of the dice score both to the models trained with FL and with HEFL. Notably, no performance drop was seen in the model trained with HEFL as compared to the model trained with conventional FL, cf. Table 1.

**Table 1:**
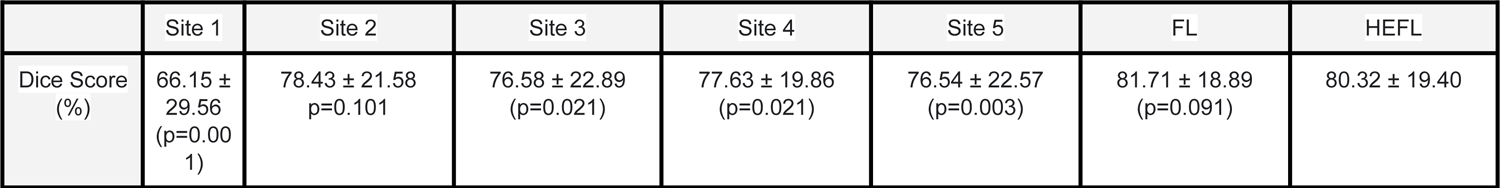
Performance of the five radiological AI models that were trained on local data only (site 1-5) and of the AI model that was trained with federated learning (FL) and with additional homomorphic encryption (HEFL). P-values are given for the comparison to HEFL.

|  | Site 1 | Site 2 | Site 3 | Site 4 | Site 5 | FL | HEFL |
| --- | --- | --- | --- | --- | --- | --- | --- |
| Dice Score (%) | 66.15 ± 29.56<br>(p=0.001) | 78.43 ± 21.58<br>p=0.101 | 76.58 ± 22.89<br>(p=0.021) | 77.63 ± 19.86<br>(p=0.021) | 76.54 ± 22.57<br>(p=0.003) | 81.71 ± 18.89<br>(p=0.091) | 80.32 ± 19.40 |

### Prediction of genetic biomarkers in colorectal cancer patients from pathology images

In an analogous setting to the radiological use-case, we tested whether HEFL performs equal to conventional FL and superior to locally trained models in the benchmarking task of predicting a molecular biomarker in colorectal cancer from pathology images: microsatellite instability (MSI)/mismatch repair deficiency (dMMR), which qualifies metastatic patients to receive cancer immunotherapy.

We performed the evaluation on independent test sets never seen during training: the clinical trial cohort QUASAR (n = 1,774 patients from the United Kingdom) and the population-based cohort YCR BCIP (Yorkshire Cancer Research Bowel Cancer Improvement Programme, n=889 patients). We trained three models on the Epi700 data (United Kingdom, n=607), the DACHS data (Germany, n=2,039) and the TCGA data (USA, n=426) respectively. Subsequently, we trained one model each in the federated learning setup including all three datasets without and with homomorphic encryption. Training with HEFL was superior to training just with local data and non-inferior to training with FL both for testing on the YCR cohort and for testing on the QUASAR cohort. Both FL and HEFL performed on the same level with no detectable difference, cf. Table 2.

**Table 2:** Area under the receiver-operator-curve for the histopathological AI models that were trained for MSI detection on the Epi700, DACHS and TCGA datasets respectively and tested on the independent QUASAR and YCR BCIP cohorts. P-values are given for the comparison to HEFL.

|  | Train on Epi700 | Train on DACHS | Train on TCGA | FL | HEFL |
| --- | --- | --- | --- | --- | --- |
| Testing on QUASAR | 74.66 ± 1.50<br>(p=0.008) | 70.38 ± 1.22<br>(p=0.001) | 70.94 ± 1.68<br>(p=0.001) | 78.52 ± 1.34<br>(p=0.289) | 79.54 ± 1.45 |
| Testing on YCR BCIP | 77.13 ± 1.74<br>(p=0.001) | 82.46 ± 2.08<br>(p=0.054) | 78.83 ± 1.67<br>(p=0.001) | 85.42 ± 1.63<br>(p=0.270) | 86.77 ± 1.65 |

### Secure training is time-efficient

A notable drawback of homomorphic encryption is its computational overhead. In our study, we eschewed this drawback by encrypting not the entire training process, but only the privacy-critical weight aggregation step, which is performed by a (potentially untrusted third party), thus enabling substantial computational savings. To determine the effect of our scheme on training time compared to FL without encryption, we conducted the following experiments on a typical hardware setup used in machine learning. As a side note, de- and encryption as well as weight aggregation is usually conducted on the central processing unit (CPU), while backpropagation during training of the networks depends on the graphics processing unit (GPU).

We found that the time required for encryption was almost negligible compared to the time required to perform the backpropagation steps and the application of weight updates: for the radiological use-case described above, less than 1% of computational time was spent on decryption, encryption and homomorphic aggregation of the weights (Figure 3d). For the histopathological use-case, less than 5% of time was used for decryption and encryption (which happens at edge) and homomorphic aggregation of the weights (which happens at the central server, Figure 3b). This difference is due to the different network architectures and different number of parameters: the histopathological use-case employs a fixed backbone feature extractor^25^ and thus has fewer parameters to optimize. Encryption and decryption scales approximately linear with the number of weights to be updated, while neural network training complexity scales more than linearly in our setup. Thus, more complex networks, such as the one used to segment brain tumors invest more computational resources in the backpropagation algorithm relative to the encryption algorithm. This is encouraging, since the relationship between training time and aggregation time is in favor of more complex networks that are usually employed when working with big data.

**Fig. 3:**
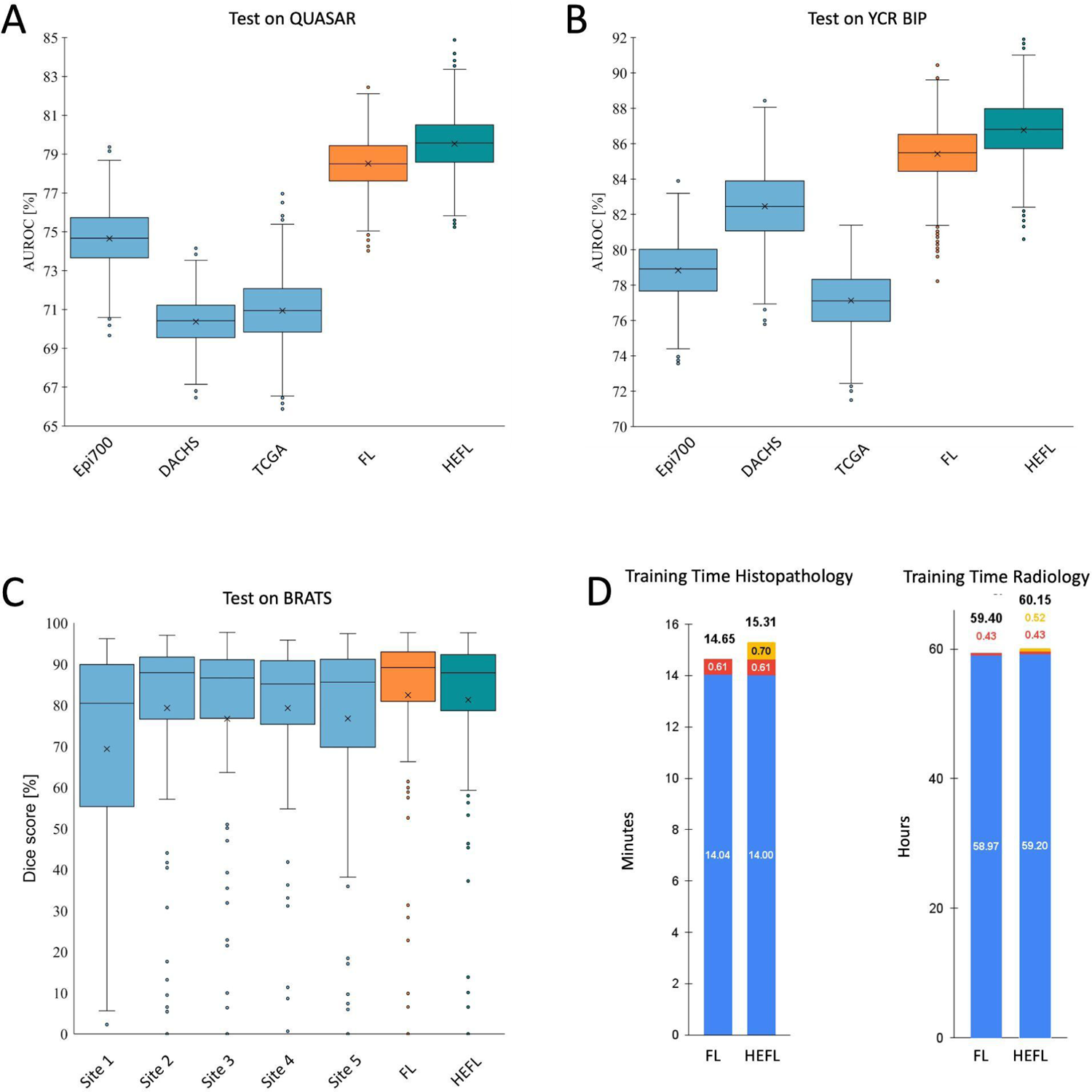
Results of training on local data only vs. training using FL and HEFL. Training neural networks on single-site datasets results in inferior performance as compared to FL and HEFL. A neural network was trained to detect MSI on data from the Epi700, the DACHS and the TCGA cohorts respectively as well as on all three datasets using FL and HEFL. The resulting networks were then tested on the QUASAR **(A)** and the YCR BIP **(B)** cohorts demonstrating superior performance of FL and HEFL. Similarly, tumor segmentation in MRI data was trained on data from five different sites as well as on all data using FL and HEFL. The resulting neural networks were then tested on an independent held-out test set and demonstrated improved performance **(C)**. Computational overhead was almost negligible (red: overhead for FL, yellow: additional overhead for encryption) as compared to training time needed for backpropagation (blue), **(D)**.

## Discussion

AI has an indisputable potential in the field of oncology ^26^ and AI models are currently reaching a stage in which they can improve patient care and render medical processes more efficient ^27, 28^. However, this improvement critically depends on the availability of sufficiently large, curated, and representative training data^29^. Currently, most research groups and industry have limited and only local data access. To train useful and generalizable AI models, stakeholders need to be able to collaborate on a large scale without jeopardizing patient privacy^26^. Only through such multi-institutional collaboration can robust AI models be trained that adequately capture the entire human population and that make the transition from bench to bedside^26^. Federated learning was initially proposed as a technical solution for privacy-preserving distributed AI^30^. FL enables joint training of AI models by multiple partners who do not share their data with each other and has been demonstrated to facilitate the training of AI models on big data^31^. Similarly, swarm learning (SL) utilizes a network of nodes to jointly train a model on distributed data and to aggregate model weights without a central instance^25^. However, FL and SL have an important shortcoming: during training, weight updates must be shared and information about the underlying data can be extracted from these weight updates as shown in our study. Such techniques should thus not be considered privacy techniques, but techniques for preserving data governance^32^. Since medical data is highly sensitive and since data privacy laws forbid the use of data in such environments, where private data can be extracted, this critically limits the applicability of collaborative learning schemes and prevents the development of powerful AI models in cancer diagnosis and treatment.

This shortcoming can be remedied by employing techniques which guarantee privacy to data owners. The only technique to guarantee privacy in a data release process is Differential Privacy^33^. Hence, when sharing the model with untrusted third parties, such a technique would have to be employed to constrain the success of attacks against patient privacy. We operate under a slightly different threat model. As all participants of the federated learning workflow described above are mutually trusting, are not intending to publish the model to the outside world and all receive an identical copy of the final model, we need only protect against an attack by the (untrusted aggregation server). Our homomorphic encryption scheme protects the weights during this critical aggregation step: local sites encrypt their weight updates before sending them out and keep the decryption key private. The entity which receives the weight updates from all sites and which performs the weight aggregation in the encrypted space thus has no access to the underlying data and no sensitive data can be extracted by design. Our technique has two notable benefits: it sidesteps the computational overhead of having to train the entire model in the encrypted space using HE. In principle, it would also be possible to use HE on all levels of the training process - i.e., also during backpropagation. However, with concurrently available computational resources, this has proven to be prohibitively computationally expensive and is not yet in reach^34^. Furthermore, as long as all data stays on site - as is the case in our FL setup - there is no need to encrypt the backpropagation procedure: potential eavesdroppers do not have access to that part of the training procedure as it is done behind secure firewalls. By restricting the fully homomorphic encryption to the most critical part of FL - the weight aggregation - we show that additional computational overload is almost negligible. Moreover, our technique allows us to avoid the privacy-utility trade-offs of employing Differential Privacy for training, in which training with Differential Privacy can lead to less-performant AI models ^20^. We note that the utilization of Differential Privacy would be mandatory in threat models different from ours, especially if the final model was designed to be shared with untrusted third parties.

A similar scheme to ours was demonstrated by Kaissis et al. in a proof-of-concept study for classifying pneumonia on chest radiographs by using secure multi-party computation through additive secret sharing^1634^. With our study, we are the first to comprehensively assess fully homomorphic encryption in cancer diagnosis on large multi-centric databases spanning both radiology and histopathology.

Our study demonstrates that AI models for oncological image-processing can be trained securely on multi-institutional data without compromising patient privacy. This will facilitate collaboration between researchers and industry alike, ultimately leading to the development of advanced and clinically useful AI models. We show that implementing the FL scheme together with homomorphic encryption comes with minimal additional code complexity and can be performed with our publicly available code.

A technical limitation of our study is that we performed all experiments within one institutional network. However, by distributing the datasets to different computing entities and keeping them strictly separate, we simulated the setting in which multiple institutions - each with their own network - perform FL realistically. We assumed a constant network communication cost in our experiments. In realistic settings, communication overhead can be unpredictable, as it depends on more factors than network size (such as concurrent traffic or physical distance of the sites). We thus chose to exclude this factor, believing it to only represent a minor limitation. We note that homomorphically encrypted weights cannot be efficiently reduced in size by compression, however this limitation is negligible compared to the requirement to encode them as 64-bit data types for transmission over HTTP. Moreover, as all parties are mutually trusting and receive an identical copy of the fully trained model at the end of training, we utilized the same key pair to encrypt the weights on all participating nodes, thus avoiding the technical challenge of key distribution.

Further improvements to the FL process are possible: with increasing peer numbers who participate in the FL setup, participation of a bounded number of malicious participants who try to corrupt the training process by delivering adversarial weight updates is possible, whereas we regarded all participants as either fully trusted or honest but curious. It has been shown that regular FL fails to converge in the presence of faulty and malicious clients^35^. Measures to counter these attacks are available and can be integrated in federated learning schemes should the need arise ^36^.

In conclusion, our study provides a blueprint for the secure and privacy-preserving multi-institutional training of oncological AI models and solves an urgent need, since it is becoming increasingly clear that differences in race and gender affect disease risk among individuals and that existing datasets at local institutions are insufficient to account for these effects.

## Methods

### Ethics statement

This study was carried out in accordance with the Declaration of Helsinki. This study is a retrospective analysis of publicly available anonymized MRI examinations and of anonymized histopathological tissue samples from multiple cohorts of cancer patients. Collection and anonymization of patients in all cohorts took place in each contributing center. Approval by the local ethics committee at each contributing center was given if applicable (QUASAR: North East – York Research Ethics Committee; YCR: Ethical approval was not required, because screening was recommended in all patients diagnosed with CRC. Testing was considered part of the ‘standard of care’ clinical pathway; Epi700: Northern Ireland Biobank (NIB13/0069, NIB13/0087, NIB13/0088 and NIB15/0168), DACHS: Ethics committee of the Medical Faculty, University of Heidelberg). Approval of the ethics committee at the University Hospital of Aachen was given for the retrospective analysis of anonymized image data under reference number “Ethikkommission EK 028/19”.

### Patient cohorts

MRI data for the BraTS patient collective contains brain MRI scans of 341 patients collected from 17 imaging centers and additional 28 patients for whom the imaging centers were not specified by the data provider. During federated learning we allocated the patients to five data clusters simulating the situation in which a regional hospital’s image database contains MRI data of multiple imaging centers. This situation is typical in real-world scenarios where patients are referred for surgery and bring their image data that had been acquired at an external institution before. The allocation of patients is detailed in supplemental table 1. All MRI examinations contained pre- and post-contrast T1-weighted sequences, T2-weighted sequences and fluid attenuation inversion-recovery sequences (FLAIR). All sequences were acquired in axial orientation. All the imaging datasets have been segmented manually, by one to four raters, following the same annotation protocol, and their annotations were approved by experienced neuro-radiologists.

For the histopathological data we collected digital whole slide images (WSI) of H&E-stained slides of human colorectal cancer (CRC) from five patient cohorts, three of which were used as training cohorts and two of which were used as test cohorts following the division of data in a previous study^25^. The training cohorts are representative of real-world clinical settings. First, the Northern Ireland Epi700 (n=661) cohort study contained data of patients with stage II and III colon cancer. This data was provided by the Northern Ireland Biobank ^37, 38^ (application NIB20-0346). Second, the “Darmkrebs: Chancen der Verhütung durch Screening” study (DACHS, n=2448) is a large population-based case-control study. This data includes samples of CRC patients at any disease stage. This data was collected from over 20 hospitals in Germany. Data collection was coordinated by the German Cancer Research Center (DKFZ, Heidelberg, Germany)^39–41^ and supported by the NCT tissue bank at the National Center for Tumor Diseases and the Institute of Pathology at the University of Heidelberg. Third, “The Cancer Genome Atlas” (TCGA) CRC cohort (n=632) is a large collection of tissue specimens from multiple populations across different countries, but largely from the United States of America (USA).^42^

We employed two separate test cohorts: The “Quick and Simple and Reliable” (QUASAR) cohort was derived from a clinical trial of adjuvant therapy containing 2206 WSI, which aimed to determine survival benefit from adjuvant chemotherapy in CRC patients from the United Kingdom (UK) ^43, 44^. The second test cohort used data from the Yorkshire Cancer Research Bowel Cancer Improvement Programme^45^ (YCR-BCIP) cohort (n=889). This was a population-based study collected in the Yorkshire Region in the UK. For all cohorts, microsatellite instability (MSI) / mismatch repair deficiency (dMMR)^46^ data were acquired.

The distribution of tumor stages in TCGA, DACHS and YCR-BCIP is comparable, see supplemental table 2. In QUASAR, stage III tumors are overrepresented due to the fact that adjuvant therapy is mainly performed in intermediate stage tumors. Therefore, following previous work^25^, we used YCR-BCIP and QUASAR as test cohorts to investigate the robustness of the AI models both on a general population and on a clinical trial population. Importantly, neither in the MRI data nor in the histopathological data, there was any overlap between training and test cohorts.

### Deep learning training procedure

The hardware used in our experiments were Intel CPUs with 18 cores and 32 GB RAM and Nvidia RTX 6000 GPUs with 24 GB memory.

#### MRI data

All of the 3D volumes were cropped around the brain to lower the computational costs and standardize the field of view. As intensity distributions vary across magnetic resonance images, intensity normalization is crucial. Therefore, we clipped the intensity values above the 99 percentile of the image, then subtracted the minimum value of the result from voxel values and divided the shifted image by the maximum value of the image. We performed data augmentation during training by applying random cropping of patches of *128 × 128 × 128* from each original volume around its center. Additionally, we applied medio-lateral and cranio-caudal flipping with a probability of *0.4*. Intensity was randomly rescaled according to a power-law *I_new_ = g. I ^γ47^* with gain *g* and the exponent γ randomly selected between *0.8 - 1.2* from a uniform distribution. White Gaussian noise with zero mean and a standard deviation of *0.03* was added to each sequence of the multi modal MRI data. A modified 4-level 3D U-Net ^48, 49^ was utilized for segmentation of brain tumors. In the contraction path, each layer contained two *3 × 3 × 3* convolutions, each followed by a rectified linear unit (ReLU), a batch normalization (BN) and then a *2 × 2 × 2* max pooling with strides of two in each dimension. The output channel number was doubled after each level in the contraction path, and it was 48 at the end of the level one. In the expansion path, each layer consisted of a nearest neighbor up-sampling of *2 × 2 × 2* in each dimension, followed by two *3 × 3 × 3* convolutions each followed by a ReLU and BN. The output channel number was halved after each level in the expansion path. In the last layer, a *1 × 1 × 1* convolution, which reduced the number of output channels to 3, followed by a SoftMax layer, was used for the per-voxel final classification.

The model was optimized using the Adam optimizer with a learning rate of 10^−4^. To be consistent in our comparison scenarios, all the weight and bias parameters of all the different models were initialized using the He initialization scheme ^50^. As a loss function, we chose the Dice loss tailored to the BraTS data needs ^51^. To minimize the overhead and make maximum use of the graphics processing unit memory, we utilized large input tiles over a large batch size and reduced the batch to a single 3D image ^49^ with 4 channels, each channel being one of the MR modalities. Hence, the batch normalization acted like instance normalization in our implementation. The network contained a total of 5,670,579 trainable parameters.

#### Histopathological data

For prediction of molecular features from image data, we based our analysis on a well established weakly-supervised end-to-end prediction pipeline, which was described and evaluated in a recent benchmark study.^52^. As a preprocessing step, the original gigapixel WSIs were tessellated into patches of size (512 × 512 × 3) pixels and were color-normalized with the Macenko method.^53^

Blurry patches and patches with no tissue were removed from the data set using canny edge detection ^52^. Following that approach, we obtained a normalized edge image using the “canny” method in Python’s OpenCV package and then removed all tiles with a mean value below a threshold of 4. A pre-trained ResNet18 was used to extract a (512 × 1) feature vector from 150 randomly selected patches for each patient ^9^. Before training, the number of tiles in each class were equalized by random undersampling until all classes had the same number of tiles, as described before ^9, 12^. Feature vectors served as input to a fully connected classification network and the patient-wise MSI label was used to label every single tile derived from that patient. The fully connected classifier network comprised four layers with (512×256), (256×256), (256×128) and (128×2) connections with a ReLU activation function.

### Code availability

Our source code for secure federated learning using homomorphic encryption is publicly available at https://github.com/tayebiarasteh/federated_HE and will be permanently archived on zenodo upon publishing of the paper. All source codes for training and evaluation of the deep neural networks, MR image analysis and preprocessing, 3D data augmentation, and gradient inversion attack are available at https://github.com/tayebiarasteh/federated_HE. All source code for the histological image analysis is available at https://github.com/KatherLab/HIA and all source code for histological image preprocessing is available at https://github.com/KatherLab/preProcessing. All code for the experiments was developed in Python 3.8 using the PyTorch 1.4 framework. The secure federated learning process was developed using PySyft 0.2.9.

### Data availability

The data that support the findings of this study are in part publicly available, in part proprietary datasets provided under collaboration agreements. Data from the BraTS collective is publicly available under https://www.med.upenn.edu/cbica/brats2020/data.html. Data (including histological images) from the TCGA database are available at https://portal.gdc.cancer.gov/. All molecular data for patients in the TCGA cohorts are available at https://cbioportal.org. Data access for the Northern Ireland Biobank can be requested at http://www.nibiobank.org/for-researchers. All other data can be requested from the respective study groups who independently manage data access for their study cohorts.

## Additional information

## Acknowledgements

The authors are grateful for the support by NVIDIA who provided counsel and supported our group with two RTX6000 GPUs. We additionally acknowledge support by the tissue bank of the National Center for Tumor Diseases (NCT) at the Institute of Pathology at University Hospital Heidelberg, Heidelberg, Germany, for providing access to the biobank data.

## Author contributions

JNK, DT and STA designed the study; STA, OLS, DT and JNK developed the software; STA performed the experiments; STA and DT and JNK analyzed the data; STA performed statistical analyses; all authors provided clinical expertise and contributed to the interpretation of the results. STA, DT and JNK wrote the manuscript, all authors corrected the manuscript and collectively made the decision to submit for publication.

## Supplemental Information

**Table S1:** Allocation of patients in the BraTS collective to five data clusters used in the federated learning setup simulating the situation in which five regional hospitals’ image databases contain a multitude of examinations from different scanners.

| Data source | number of examinations | Allocation to Federated Learning Center # |
| --- | --- | --- |
| BraTS center 1 | 129 | 1 |
| BraTS center 2 | 14 | 2 |
| BraTS center 3 | 5 | 2 |
| BraTS center 4 | 9 | 2 |
| BraTS center 5 | 22 | 2 |
| BraTS center 6 | 34 | 3 |
| BraTS center 7 | 12 | 3 |
| BraTS center 8 | 8 | 3 |
| BraTS center 9 | 4 | 3 |
| BraTS center 10 | 8 | 4 |
| BraTS center 11 | 6 | 4 |
| BraTS center 12 | 9 | 4 |
| BraTS center 13 | 27 | 4 |
| BraTS center 14 | 3 | 4 |
| BraTS center 15 | 11 | 4 |
| BraTS center 16 | 34 | 5 |
| BraTS center 17 | 6 | 5 |
| BraTS unknown group | 28 | 5 |
| Sum | 369 |  |

**Table S2:** Clinico-pathological features of histopathological cohorts in our study. Abbreviations: colorectal cancer (CRC), number of patients (N Patients), interquartile range (IQR), microsatellite instability (MSI), microsatellite stability (MSS). Definitions: Right-sided CRC from cecum to transverse colon.

|  | TCGA | DACHS | Epi700 | QUASAR | YCR-BCIP |
| --- | --- | --- | --- | --- | --- |
| N Patients | 632 | 2448 | 661 | 2190 | 889 |
| Age (median ± IQR) | 68 ± 18 | 69 ± 14 | 72 ± 14 | 63 ± 12 | 71 ± 15 |
| Gender: Male | 322 (50.9%) | 1436 (58.7%) | 358 (54.2%) | 1334 (60.9%) | 494 (55.6%) |
| Gender: Female | 292 (46.2%) | 1012 (41.3%) | 303 (45.8%) | 848 (38.7%) | 395 (44.4%) |
| Gender: Unknown | 18 (2.85%) | 0 | 0 | 8 (0.4%) | 0 |
| MSS/pMMR | 392 (62%) | 1836 (75%) | 471 (71.3%) | 1529 (69.8%) | 760 (85.5%) |
| MSI/dMMR | 65 (10.3%) | 210 (8.6%) | 136 (20.6%) | 246 (11.2%) | 129 (14.5%) |
| unknown MSI status | 175 (27.7%) | 402 (16.4%) | 54 (8.1%) | 415 (19%) | 0 |
| Stage 1 | 76 (12%) | 485 (19.8%) | 0 | 5 (0.2%) | 169 (19%) |
| Stage 2 | 166 (26.3%) | 801 (32.7%) | 394 (59.6%) | 53 (2.4%) | 317 (35.7%) |
| Stage 3 | 140 (22.2%) | 822 (33.6%) | 267 (40.4%) | 1653 (75.5%) | 370 (41.6%) |
| Stage 4 | 63 (10%) | 337 (13.8%) | 0 (0%) | 268 (12.2%) | 33 (3.7%) |
| Stage unknown | 187 (29.5) | 3 (0.1%) | 0 | 211 (9.7%) | 0 |
| Left-sided CRC | 248 (39.2%) | 1607 (65.6%) | 280 (42.3%) | 1158 (52.9%) | 487 (54.8%) |
| Right-sided CRC | 176 (27.8%) | 819 (33.5%) | 375 (56.7%) | 754 (34.4%) | 332 (37.3%) |
| Unknown side | 209 (33%) | 22 (0.9%) | 6 (1%) | 278 (12.7%) | 70 (7.9%) |

## References

1. Kleppe, A. et al. Designing deep learning studies in cancer diagnostics. Nat. Rev. Cancer (2021) doi:10.1038/s41568-020-00327-9.

2. Boehm, K. M., Khosravi, P., Vanguri, R., Gao, J. & Shah, S. P. Harnessing multimodal data integration to advance precision oncology. Nat. Rev. Cancer (2021) doi:10.1038/s41568-021-00408-3.

3. Echle, A. et al. Deep learning in cancer pathology: a new generation of clinical biomarkers. British Journal of Cancer (2020) doi:10.1038/s41416-020-01122-x.

4. Elemento, O., Leslie, C., Lundin, J. & Tourassi, G. Artificial intelligence in cancer research, diagnosis and therapy. Nat. Rev. Cancer (2021) doi:10.1038/s41568-021-00399-1.

5. Kather, J. N. & Calderaro, J. Development of AI-based pathology biomarkers in gastrointestinal and liver cancer. Nat. Rev. Gastroenterol. Hepatol. (2020) doi:10.1038/s41575-020-0343-3.

6. Lu, M. Y. et al. AI-based pathology predicts origins for cancers of unknown primary. Nature 594, 106–110 (2021).

7. Lotter, W. et al. Robust breast cancer detection in mammography and digital breast tomosynthesis using an annotation-efficient deep learning approach. Nat. Med. 27, 244–249 (2021).

8. Coudray, N. et al. Classification and mutation prediction from non–small cell lung cancer histopathology images using deep learning. Nat. Med. 24, 1559–1567 (2018).

9. Kather, J. N. et al. Deep learning can predict microsatellite instability directly from histology in gastrointestinal cancer. Nat. Med. 25, 1054–1056 (2019).

10. Loeffler, C. M. L. et al. Artificial Intelligence–based Detection of FGFR3 Mutational Status Directly from Routine Histology in Bladder Cancer: A Possible Preselection for Molecular Testing? European Urology Focus (2021) doi:10.1016/j.euf.2021.04.007.

11. Fu, Y. et al. Pan-cancer computational histopathology reveals mutations, tumor composition and prognosis. Nature Cancer 1–11 (2020).

12. Kather, J. N. et al. Pan-cancer image-based detection of clinically actionable genetic alterations. Nature Cancer 1, 789–799 (2020).

13. Binder, A. et al. Morphological and molecular breast cancer profiling through explainable machine learning. Nature Machine Intelligence 3, 355–366 (2021).

14. McMahan, B., Moore, E., Ramage, D., Hampson, S. & Arcas, B. A. y. Communication-Efficient Learning of Deep Networks from Decentralized Data. in Proceedings of the 20th International Conference on Artificial Intelligence and Statistics (eds. Singh, A. & Zhu, J.) vol. 54 1273–1282 (PMLR, 2017).

15. Lu, M. Y., et al. Federated Learning for Computational Pathology on Gigapixel Whole Slide Images. arXiv [eess.IV] (2020).

16. Kaissis, G. et al. End-to-end privacy preserving deep learning on multi-institutional medical imaging. Nature Machine Intelligence 3, 473–484 (2021).

17. Wang, Z., Song, M., Zhang, Z. & Song, Y. Beyond inferring class representatives: User-level privacy leakage from federated learning. IEEE INFOCOM 2019 (2019).

18. Kaissis, G. A., Makowski, M. R., Rückert, D. & Braren, R. F. Secure, privacy-preserving and federated machine learning in medical imaging. Nature Machine Intelligence 2, 305–311 (2020).

19. Usynin, D. et al. Adversarial interference and its mitigations in privacy-preserving collaborative machine learning. Nature Machine Intelligence 3, 749–758 (2021).

20. Lu, M. Y. et al. Federated learning for computational pathology on gigapixel whole slide images. Med. Image Anal. 76, 102298 (2022).

21. Menze, B. H. et al. The Multimodal Brain Tumor Image Segmentation Benchmark (BRATS). IEEE Trans. Med. Imaging 34, 1993–2024 (2015).

22. Bakas, S. et al. Advancing The Cancer Genome Atlas glioma MRI collections with expert segmentation labels and radiomic features. Sci Data 4, 170117 (2017).

23. Bakas, S., et al. Identifying the Best Machine Learning Algorithms for Brain Tumor Segmentation, Progression Assessment, and Overall Survival Prediction in the BRATS Challenge. arXiv [cs.CV] (2018).

24. Zhao, B., Mopuri, K. R. & Bilen, H. iDLG: Improved Deep Leakage from Gradients. arXiv [cs.LG] (2020).

25. Saldanha, O. L., et al. Swarm learning for decentralized artificial intelligence in cancer histopathology. (2021).

26. Bhinder, B., Gilvary, C., Madhukar, N. S. & Elemento, O. Artificial Intelligence in Cancer Research and Precision Medicine. Cancer Discov. 11, 900–915 (2021).

27. Killock, D. AI outperforms radiologists in mammographic screening. Nature reviews. Clinical oncology vol. 17 134 (2020).

28. McKinney, S. M. et al. International evaluation of an AI system for breast cancer screening. Nature 577, 89–94 (2020).

29. Willemink, M. J. et al. Preparing Medical Imaging Data for Machine Learning. Radiology 295, 4–15 (2020).

30. Konečný, J., et al. Federated Learning: Strategies for Improving Communication Efficiency. *arXiv [cs.LG]* (2016).

31. Dayan, I. et al. Federated learning for predicting clinical outcomes in patients with COVID-19. Nat. Med. 27, 1735–1743 (2021).

32. Ziller, A., Mueller, T. T., Braren, R., Rueckert, D. & Kaissis, G. Privacy: An Axiomatic Approach. Entropy 24, (2022).

33. Dwork, C. & Roth, A. The Algorithmic Foundations of Differential Privacy. Found. Trends Theor. Comput. Sci. 9, 211–407 (2014).

34. Keller, M., Pastro, V. & Rotaru, D. Overdrive: Making SPDZ Great Again. in Advances in Cryptology – EUROCRYPT 2018 158–189 (Springer International Publishing, 2018).

35. Blanchard, P., El Mhamdi, E. M., Guerraoui, R. & Stainer, J. Machine learning with adversaries: Byzantine tolerant gradient descent. Adv. Neural Inf. Process. Syst. 30, (2017).

36. Ma, X., Zhou, Y., Wang, L. & Miao, M. Privacy-preserving Byzantine-robust federated learning. Comput. Stand. Interfaces 80, 103561 (2022).

37. Lewis, C. et al. The northern Ireland biobank: A cancer focused repository of science. Open J. Bioresour. 5, (2018).

38. Loughrey, M. B. et al. Identifying mismatch repair-deficient colon cancer: near-perfect concordance between immunohistochemistry and microsatellite instability testing in a large, population-based series. Histopathology 78, 401–413 (2021).

39. Carr, P. R. et al. Estimation of Absolute Risk of Colorectal Cancer Based on Healthy Lifestyle, Genetic Risk, and Colonoscopy Status in a Population-Based Study. Gastroenterology vol. 159 129–138.e9 (2020).

40. Brenner, H., Chang-Claude, J., Seiler, C. M., Stürmer, T. & Hoffmeister, M. Does a negative screening colonoscopy ever need to be repeated? Gut 55, 1145–1150 (2006).

41. Li, X., Jansen, L., Chang-Claude, J., Hoffmeister, M. & Brenner, H. Risk of Colorectal Cancer Associated With Lifetime Excess Weight. JAMA Oncol 8, 730–737 (2022).

42. GDC. https://portal.gdc.cancer.gov.

43. QUASAR Collaborative Group. Adjuvant chemotherapy versus observation in patients with colorectal cancer: a randomised study. Lancet 370, 2020–2029 (2007).

44. Hutchins, G. et al. Value of mismatch repair, KRAS, and BRAF mutations in predicting recurrence and benefits from chemotherapy in colorectal cancer. J. Clin. Oncol. 29, 1261–1270 (2011).

45. Taylor, J. et al. Regional multidisciplinary team intervention programme to improve colorectal cancer outcomes: study protocol for the Yorkshire Cancer Research Bowel Cancer Improvement Programme (YCR BCIP). BMJ Open vol. 9 e030618 (2019).

46. Marks, K. & West, N. Molecular assessment of colorectal cancer through Lynch syndrome screening. Diagn. Histopathol. 26, 47–50 (2020).

47. Cirillo, M. D., Abramian, D. & Eklund, A. What is the best data augmentation for 3D brain tumor segmentation? arXiv [eess.IV] (2020).

48. Çiçek, Ö., Abdulkadir, A., Lienkamp, S. S. & Brox, T. 3D U-Net: learning dense volumetric segmentation from sparse annotation. conference on medical … (2016).

49. Ronneberger, O., Fischer, P. & Brox, T. U-net: Convolutional networks for biomedical image segmentation. Med. Image Comput. Comput. Assist. Interv. (2015).

50. He, K., Zhang, X., Ren, S. & Sun, J. Delving deep into rectifiers: Surpassing human-level performance on imagenet classification. Proc. IEEE (2015).

51. Henry, T., et al. Brain Tumor Segmentation with Self-ensembled, Deeply-Supervised 3D U-Net Neural Networks: A BraTS 2020 Challenge Solution. in Brainlesion: Glioma, Multiple Sclerosis, Stroke and Traumatic Brain Injuries 327–339 (Springer International Publishing, 2021).

52. Laleh, N. G. et al. Benchmarking artificial intelligence methods for end-to-end computational pathology. bioRxiv 2021.08.09.455633 (2021) doi:10.1101/2021.08.09.455633.

53. Macenko, M. et al. A method for normalizing histology slides for quantitative analysis. in 2009 IEEE International Symposium on Biomedical Imaging: From Nano to Macro 1107–1110 (2009).

